# Association between skull bone mineral density and periodontitis: evidence from the National Health and Nutrition Examination Survey (2011-2014)

**DOI:** 10.1101/2022.07.04.22277233

**Authors:** Fuqian Jin, Jukun Song, Yi Luo, Beichuan Wang, Ming Ding, Jiaxin Hu, Zhu Chen

**Affiliations:** School of Stomatology, Zunyi Medical University, Zunyi, Guizhou, China; Department of Oral Medicine, Guiyang Hospital of Stomatology, Guiyang, Guizhou, China; Department of Oral and Maxillofacial Surgery, Guizhou Provincial People’s Hospital, Guiyang, Guizhou, China

## Abstract

**Background and Objective:** Bone mineral density (BMD) and periodontitis have been the subject of many studies. However, the relationship between skull (including mandible) BMD and periodontitis has not been extensively studied. An objective of this cross-sectional study was to examine the relationship between skull BMD and periodontitis using data from the National Health and Nutrition Examination Surveys (NHANES) for 2011-2012 and 2013-2014.

**Materials and Methods:** From 19,931 participants, 3,802 were screened and included with no missing values in the study. We examined the distribution of variables by grouping the skull BMD levels into quartiles. Periodontitis is defined by the Centers for Disease Control and Prevention (CDC) and the American Periodontal Association (AAP) in 2012. An interaction test was conducted using stratified and adjusted logistic regression models, and multivariate logistic regression analysis was performed, along with curve fitting and a threshold effect analysis were performed on the relationship between skull BMD and periodontitis.

**Results:** The results showed a negatively relationship between skull BMD and the risk of periodontitis. Although the inflection point was found (the skull BMD= 2.89g/cm^2^), it was not statistically significant, indicating that the skull BMD and periodontitis are linearly related, which 1 unit increase in the skull BMD (g/cm^2^) was associated with a 30% (OR=0.70; CI=0.57, 0.87; p=0.0010) reduction in the risk of periodontitis events.

**Conclusions:** Periodontal disease may be related to low skull BMD, for those people, oral hygiene and health care should be more closely monitored. Validation of our findings will require further research.

## Introduction

The burden of periodontitis continues to be a worldwide public health problem, and the majority of periodontitis incidence is observed in those between 55 and 59 years of age, while younger people are experiencing an increasing incidence of periodontitis(1). Periodontitis is chronic inflammation of the tissue supporting teeth, and if it progresses, it can lead to alveolar bone loss and ultimately tooth loss(2). It is intended that periodontitis occurs when dental plaque accumulates on the teeth and results in an imbalance between bacterial invasion and host defense(3). Meanwhile, host responses to general health conditions as well as be associated with periodontitis(4, 5). Osteoporosis and periodontitis have been associated in most cross-sectional studies, especially for postmenopausal women(6-9).

Bone mineral density (BMD) measures are the optimal method for diagnosing osteoporosis and osteopenia. The World Health Organization recommends using dual energy X-ray absorptiometry (DXA) to assess the BMD of the spine, hip, and forearm(10). Apparently, osteoporosis has been associated with periodontal disease risk periodontal diseases(11). Human osteoporosis may negatively impact alveolar bone height, but periodontitis-induced bone loss is not affected by skeletal homeostasis (12). A study by Munhoz et al. documented the BMD of mandible was measured by DXA in systemically healthy subjects and then found that the low BMD of mandible may be related to chronic periodontitis(13). The mandible has high bone turnover, increased blood flow, and is more sensitive to osteoclast and osteoblast activity than any other sites, however, researchers encountered difficulties working with the mandible because of its complex bone mineral distribution (10). Meanwhile, the skull is connected by irregular shapes and thickness of the bones, fibrous joints, and complex muscle relationships, which seems to rule out the possibility of analyzing a single bone(14). The skull (including mandible) BMD, which as part of the whole-body BMD measurement, is well correlated to the rest of the skeleton(15). Due to very little mechanical strain and weight bearing on the skull, it is a unique part of the skeleton, at the same time, the measurement of a skull’s BMD may be used to screen for hereditary diseases, skeletal artifacts, or to assess oral bone loss (15, 16).Athletes with stress sites in their skeleton can increase bone density through impact loading sports, the study by Courteix et al.(17) reported that in gymnasts, the skull BMD is lower than other people because of the absence of stress. It is possible to screen for osteoporosis by measuring BMD with axial skull CT because patients with a positive head CT scan for the condition are twice as likely to suffer fractures as healthy people are(18).

Research on the relationship between skull BMD and periodontitis in a large and representative population is necessary to develop. Due to this, we analyzed secondary data based on available data from NHANES. Study objectives are to determine if there is a significant relationship between skull BMD and periodontitis and to understand the associated confounders.

## Materials and methods

### Data source

In the present cross-sectional retrospective analysis, continuous National Health and Nutrition Examination Survey (NHANES) data from cycles 2011-2012 and 2013-2014 were analyzed. The Centers for Disease Control and Prevention (CDC) manages the National Center for Health Statistics (NCHS) which conducts the NHANES to assess the health and nutritional status of children, adults, and the elderly. Approval of both datasets was granted by NCHS Research Ethics Review Board (ERB), protocol #2011-17 and informed consent was received from all participants. Authors did not collect the data but obtained it from the CDC website which can be downloaded for free (https://wwwn.cdc.gov/nchs/nhanes/Default.aspx). This manuscript meets the criteria stated at the STROBE guidelines.

### Study population

There were 19,931 participants in the two cycles of 2011-2012 and 2013-2014, we selected participants who integrally underwent skull BMD assessment(n=10,104), which including 8-59 years old. In total, 4,123 participants provided complete oral examination, which including 30-59 years old. Then those who simultaneous with incomplete clinical and sociodemographic data were removed. In the study, 3802 out of 19,931 participants were screened.

### Variables

The independent variable in the present study was the skull BMD (g/cm^2^), which with Apex 3.2 software, scans were acquired on the Hologic Discovery model A densitometers (Hologic, Inc, Bedford, Massachusetts) and participants aged 8-59 years were eligible. The targeted dependent variable was periodontitis (dichotomous variable). Mobile examination centers (MEC) were used to conduct the periodontal examinations of participants 30 years and older. For the oral health examination, the NHANES operating manuals described the training and calibration processes. Clinical attachment loss (AL) was defined as the distance between the cement-enamel junction (CEJ) and the sulcus base, and the probing depth (PD) was defined as the distance between the free gingival margin (FGM) and the sulcus base. Based on the 2012 CDC/American Academy of Periodontology (AAP) periodontitis case definitions, participants must have at least two teeth that meet specific probing thresholds(19). Specifically, individuals who exhibited at least two interproximal sites with an attachment loss of at least 3 mm and at least two interproximal sites with probing depths of at least 4 mm which are not on the same tooth or at least one site with a probing depth of at least 5 mm, were considered to have at least mild periodontitis. In this study, mild, moderate and severe periodontitis were uniformly classified as periodontitis group.

Covariates include demographic, examination, and questionnaire variables. Demographic variables include gender, age (30-39, 40-49, and 50–69 years), race, education (more than high school, high school and less than high school), poverty income ratio, which was divided into (PIR <1, 1≤PIR<2, PIR≥2). Examination variables include comprehensive grip strength (kg), which was divided into three equal parts (low, middle, and high). Meanwhile, body mass index (BMI) was classified into three groups (<25, 25-30, and ≥30). The questionnaire variables include the last 7 days frequency of floss use (<4 times,≥4times), whether smoking (smoked at least 100 cigarettes in life?). Drinking (Had at least 12 alcohol drinks/1yr?) Depression, using the PHQ-9, you can determine the frequency of nine depressive symptoms(20). General health condition. Whether the doctor diagnosed hypertension, diabetes and hypertriglyceridemia. We divided the sleep time by the optimal sleep time reported in the literature: <7 hours and 7 hours(21). A detailed description of the variables can be found on the NHANES website.

### Statistical analysis

Variables that are continuous in this study are converted into categorical ones, and categorical variables are expressed by frequency or percentage. Data analysis focused mainly on these three points this study: (a) which factors modifying or interfering with the relationship between skull BMD and periodontitis; (b) when the interference factors have been adjusted or after stratifying, what is the real relationship between skull BMD and periodontitis; and (c) do skull BMD and periodontitis have a linear or non-linear relationship? As a result, three main steps can be outlined for data analysis. Firstly, we examined the distribution of variables by grouping the skull BMD levels into quartiles and then, an interaction test was conducted using stratified. Secondly, multivariate logistic regression analysis was performed by constructing three statistical models: model I, no covariates are adjusted; model II, sociodemographic data are adjusted only; model III, all covariates were adjusted. Finally, a smooth curve fitting (penalized spline method) and a threshold effect analysis were performed, calculating the inflection point using a recursive procedure and constructing a two-stage weighted linear regression model on either side of the inflection point. A recursive algorithm is used to determine the inflection point, then we build a weighted two-stage linear regression model on both sides of the inflection point. Log likelihood ratio tests are used to determine whether the model is a linear regression model or two piecewise linear regression models. In order to make sure that our data analysis was robust, we carried out sensitivity analyses as follows: According to the quartile level of skull BMD, we divided the samples into four groups, moreover, we transformed the quartile classification variables into continuous variables and calculated the trend P. We examined the possibility of nonlinearity by using skull BMD as a continuous variable. All the analyses were performed with the statistical software packages R (http://www.R-project.org, The R Foundation) and Empower Stats (http://www.empowerstats.com, X&Y Solutions, Inc, Boston, MA). All tests were two-sided and P values lower than 0.05 were considered statistically significant.

## Results

### Baseline characteristics of participants

An overview of the baseline characteristics of selected participants from the NHANES 2009 to 2014 is shown in Table 1. Differences in the distribution of poverty income ratio, hours of sleep, Frequency of floss use (last 7 days), alcohol consumption, hypertriglyceridemia, depressive symptoms, general health condition in four skull BMD groups (quartiles, Q1-Q4) were not statistically significant (all p values > 0.0 5). In comparison to Q4 group, high skull BMD subjects were periodontal health participants, female, 40-49 years old, more than high school education level, BMI≥30, low combined grip strength, no smoking, no hypertension and diabetes history.

**Table 1.**
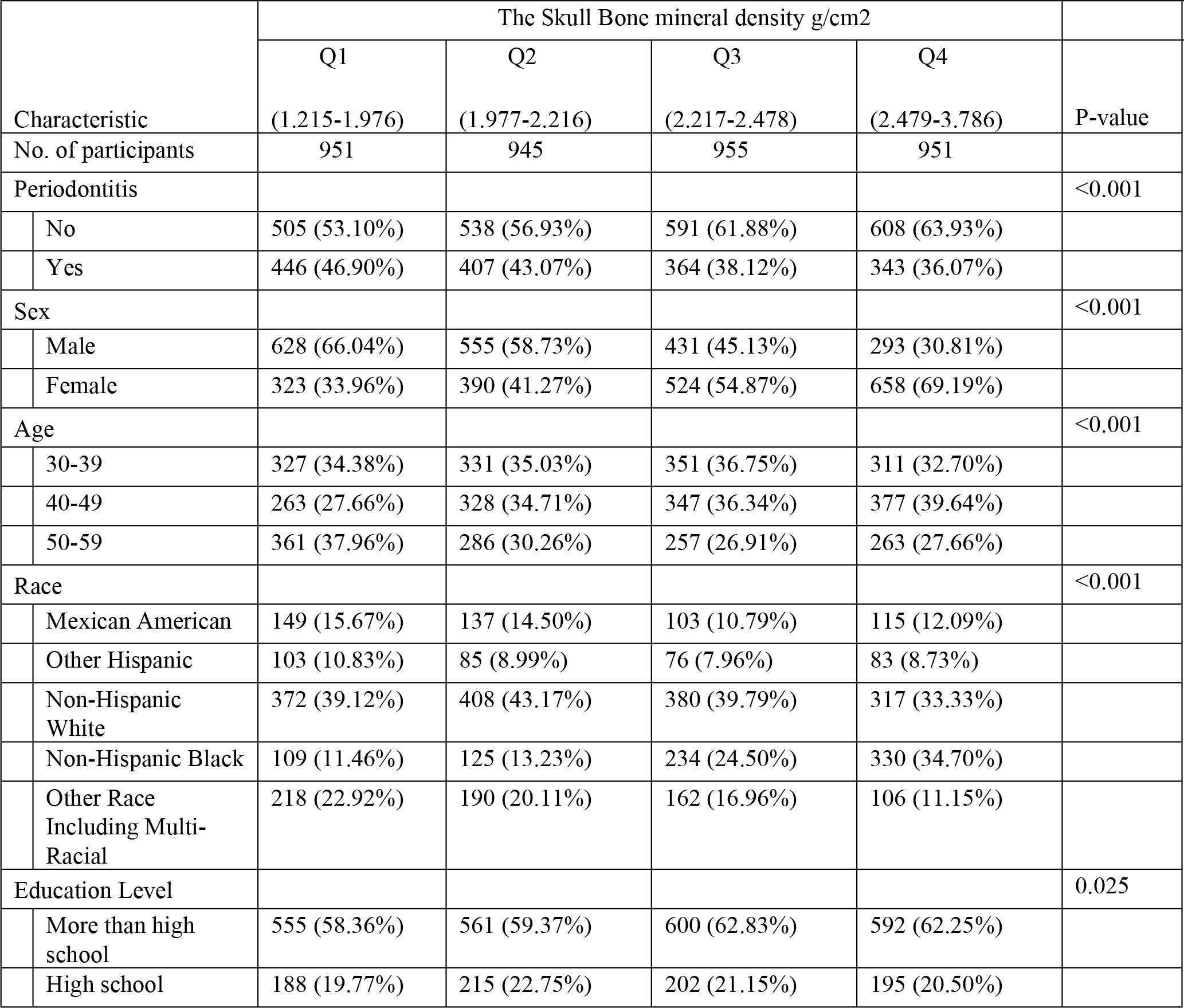
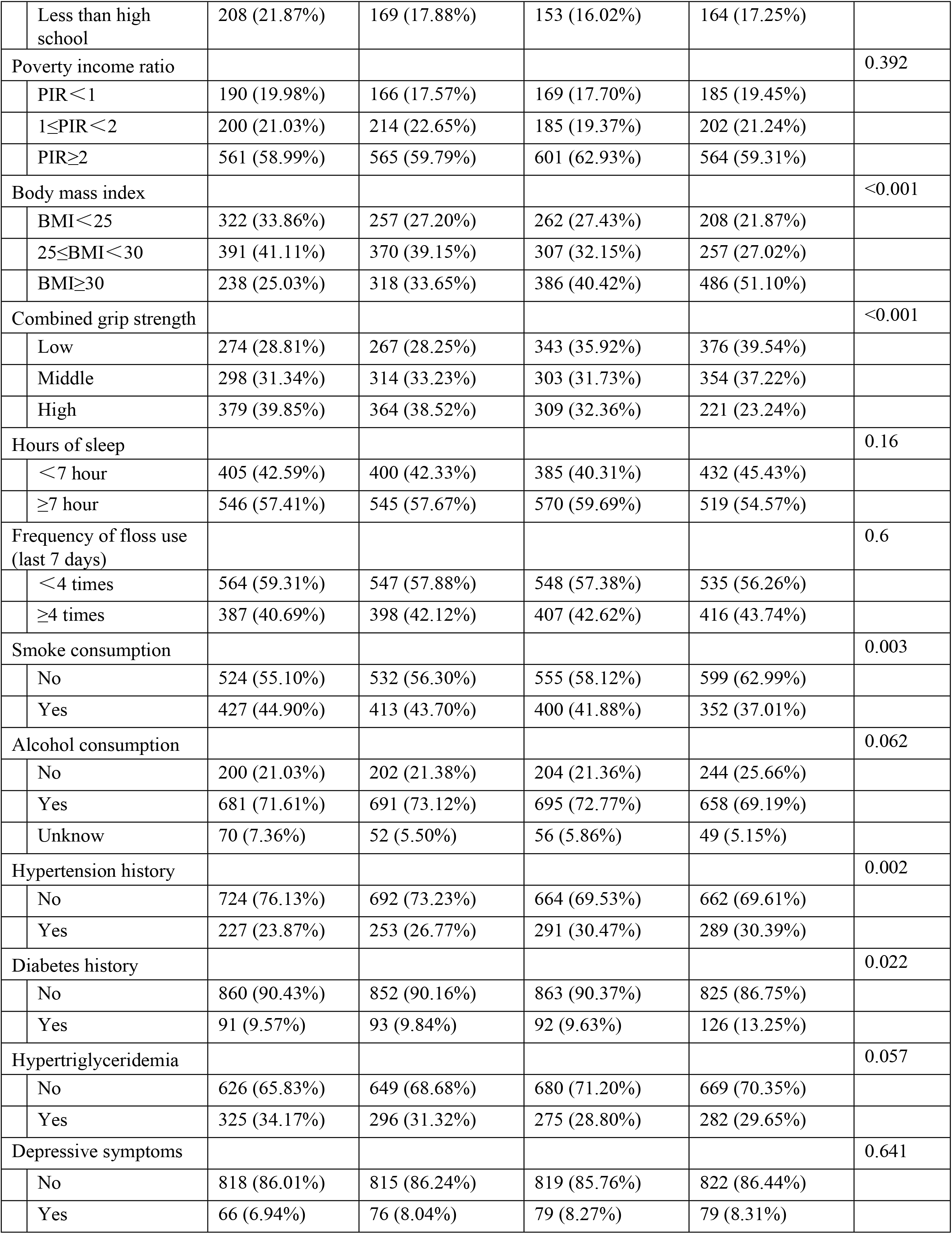
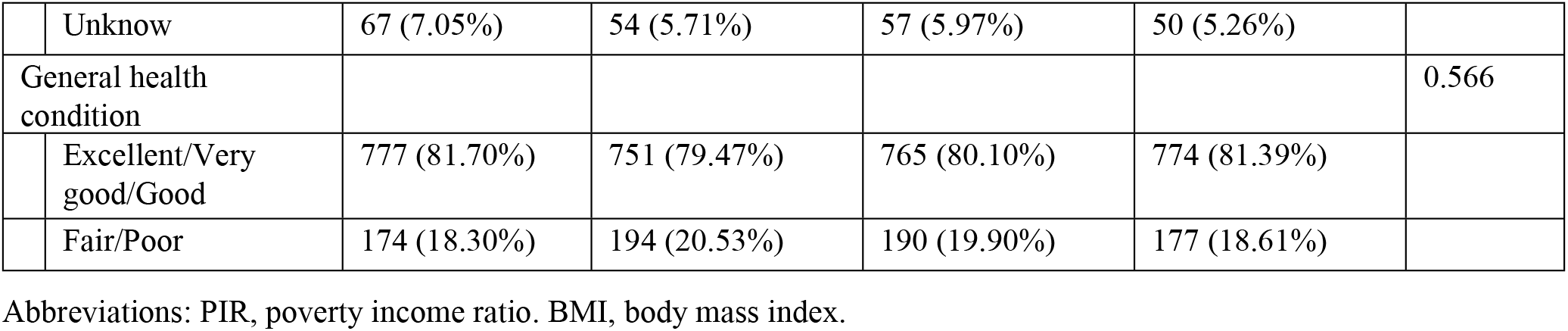
Baseline Characteristics of participants (N=3802)

| Characteristic | The Skull Bone mineral density g/cm<sup>2</sup> |  |  |  | P-value |
| --- | --- | --- | --- | --- | --- |
|  | Q1<br>(1.215-1.976) | Q2<br>(1.977-2.216) | Q3<br>(2.217-2.478) | Q4<br>(2.479-3.786) |  |
| No. of participants | 951 | 945 | 955 | 951 |  |
| Periodontitis |  |  |  |  | <0.001 |
| No | 505 (53.10%) | 538 (56.93%) | 591 (61.88%) | 608 (63.93%) |  |
| Yes | 446 (46.90%) | 407 (43.07%) | 364 (38.12%) | 343 (36.07%) |  |
| Sex |  |  |  |  | <0.001 |
| Male | 628 (66.04%) | 555 (58.73%) | 431 (45.13%) | 293 (30.81%) |  |
| Female | 323 (33.96%) | 390 (41.27%) | 524 (54.87%) | 658 (69.19%) |  |
| Age |  |  |  |  | <0.001 |
| 30-39 | 327 (34.38%) | 331 (35.03%) | 351 (36.75%) | 311 (32.70%) |  |
| 40-49 | 263 (27.66%) | 328 (34.71%) | 347 (36.34%) | 377 (39.64%) |  |
| 50-59 | 361 (37.96%) | 286 (30.26%) | 257 (26.91%) | 263 (27.66%) |  |
| Race |  |  |  |  | <0.001 |
| Mexican American | 149 (15.67%) | 137 (14.50%) | 103 (10.79%) | 115 (12.09%) |  |
| Other Hispanic | 103 (10.83%) | 85 (8.99%) | 76 (7.96%) | 83 (8.73%) |  |
| Non-Hispanic White | 372 (39.12%) | 408 (43.17%) | 380 (39.79%) | 317 (33.33%) |  |
| Non-Hispanic Black | 109 (11.46%) | 125 (13.23%) | 234 (24.50%) | 330 (34.70%) |  |
| Other Race Including Multi-Racial | 218 (22.92%) | 190 (20.11%) | 162 (16.96%) | 106 (11.15%) |  |
| Education Level |  |  |  |  | 0.025 |
| More than high school | 555 (58.36%) | 561 (59.37%) | 600 (62.83%) | 592 (62.25%) |  |
| High school | 188 (19.77%) | 215 (22.75%) | 202 (21.15%) | 195 (20.50%) |  |

|  |  |  |  |  |  |  |
| --- | --- | --- | --- | --- | --- | --- |
|  | Less than high school | 208 (21.87%) | 169 (17.88%) | 153 (16.02%) | 164 (17.25%) |  |
| Poverty income ratio |  |  |  |  |  | 0.392 |
|  | PIR < 1 | 190 (19.98%) | 166 (17.57%) | 169 (17.70%) | 185 (19.45%) |  |
|  | 1 ≤ PIR < 2 | 200 (21.03%) | 214 (22.65%) | 185 (19.37%) | 202 (21.24%) |  |
|  | PIR ≥ 2 | 561 (58.99%) | 565 (59.79%) | 601 (62.93%) | 564 (59.31%) |  |
| Body mass index |  |  |  |  |  | <0.001 |
|  | BMI < 25 | 322 (33.86%) | 257 (27.20%) | 262 (27.43%) | 208 (21.87%) |  |
|  | 25 ≤ BMI < 30 | 391 (41.11%) | 370 (39.15%) | 307 (32.15%) | 257 (27.02%) |  |
|  | BMI ≥ 30 | 238 (25.03%) | 318 (33.65%) | 386 (40.42%) | 486 (51.10%) |  |
| Combined grip strength |  |  |  |  |  | <0.001 |
|  | Low | 274 (28.81%) | 267 (28.25%) | 343 (35.92%) | 376 (39.54%) |  |
|  | Middle | 298 (31.34%) | 314 (33.23%) | 303 (31.73%) | 354 (37.22%) |  |
|  | High | 379 (39.85%) | 364 (38.52%) | 309 (32.36%) | 221 (23.24%) |  |
| Hours of sleep |  |  |  |  |  | 0.16 |
|  | < 7 hour | 405 (42.59%) | 400 (42.33%) | 385 (40.31%) | 432 (45.43%) |  |
|  | ≥ 7 hour | 546 (57.41%) | 545 (57.67%) | 570 (59.69%) | 519 (54.57%) |  |
| Frequency of floss use (last 7 days) |  |  |  |  |  | 0.6 |
|  | < 4 times | 564 (59.31%) | 547 (57.88%) | 548 (57.38%) | 535 (56.26%) |  |
|  | ≥ 4 times | 387 (40.69%) | 398 (42.12%) | 407 (42.62%) | 416 (43.74%) |  |
| Smoke consumption |  |  |  |  |  | 0.003 |
|  | No | 524 (55.10%) | 532 (56.30%) | 555 (58.12%) | 599 (62.99%) |  |
|  | Yes | 427 (44.90%) | 413 (43.70%) | 400 (41.88%) | 352 (37.01%) |  |
| Alcohol consumption |  |  |  |  |  | 0.062 |
|  | No | 200 (21.03%) | 202 (21.38%) | 204 (21.36%) | 244 (25.66%) |  |
|  | Yes | 681 (71.61%) | 691 (73.12%) | 695 (72.77%) | 658 (69.19%) |  |
|  | Unknow | 70 (7.36%) | 52 (5.50%) | 56 (5.86%) | 49 (5.15%) |  |
| Hypertension history |  |  |  |  |  | 0.002 |
|  | No | 724 (76.13%) | 692 (73.23%) | 664 (69.53%) | 662 (69.61%) |  |
|  | Yes | 227 (23.87%) | 253 (26.77%) | 291 (30.47%) | 289 (30.39%) |  |
| Diabetes history |  |  |  |  |  | 0.022 |
|  | No | 860 (90.43%) | 852 (90.16%) | 863 (90.37%) | 825 (86.75%) |  |
|  | Yes | 91 (9.57%) | 93 (9.84%) | 92 (9.63%) | 126 (13.25%) |  |
| Hypertriglyceridemia |  |  |  |  |  | 0.057 |
|  | No | 626 (65.83%) | 649 (68.68%) | 680 (71.20%) | 669 (70.35%) |  |
|  | Yes | 325 (34.17%) | 296 (31.32%) | 275 (28.80%) | 282 (29.65%) |  |
| Depressive symptoms |  |  |  |  |  | 0.641 |
|  | No | 818 (86.01%) | 815 (86.24%) | 819 (85.76%) | 822 (86.44%) |  |
|  | Yes | 66 (6.94%) | 76 (8.04%) | 79 (8.27%) | 79 (8.31%) |  |
|  | Unknow | 67 (7.05%) | 54 (5.71%) | 57 (5.97%) | 50 (5.26%) |  |
| General health condition |  |  |  |  |  | 0.566 |
|  | Excellent/Very good/Good | 777 (81.70%) | 751 (79.47%) | 765 (80.10%) | 774 (81.39%) |  |
|  | Fair/Poor | 174 (18.30%) | 194 (20.53%) | 190 (19.90%) | 177 (18.61%) |  |

Abbreviations: PIR, poverty income ratio. BMI, body mass index.

### Subgroup analyses

In Table 2, the results of a subgroup analysis examining the associations between skull BMD and periodontitis are presented. A negative correlation was found between skull BMD and periodontitis, and alcohol consumption (No: OR = 0.96 95%CI: 0.63-1.46; Yes: OR = 0.66, 95%CI: 0.51-0.86, P for interaction=0.023) was associated with it even more. The effect of the skull BMD on periodontitis showed no difference in the following subgroups: sex, age, race, education levels, poverty income ratio, body mass index, combined grip strength, hours of sleep, Frequency of floss use (last 7 days), smoke consumption, hypertension history, diabetes history, and general health condition (all P for interaction >0.10) (Table 2)

**Table 2.**
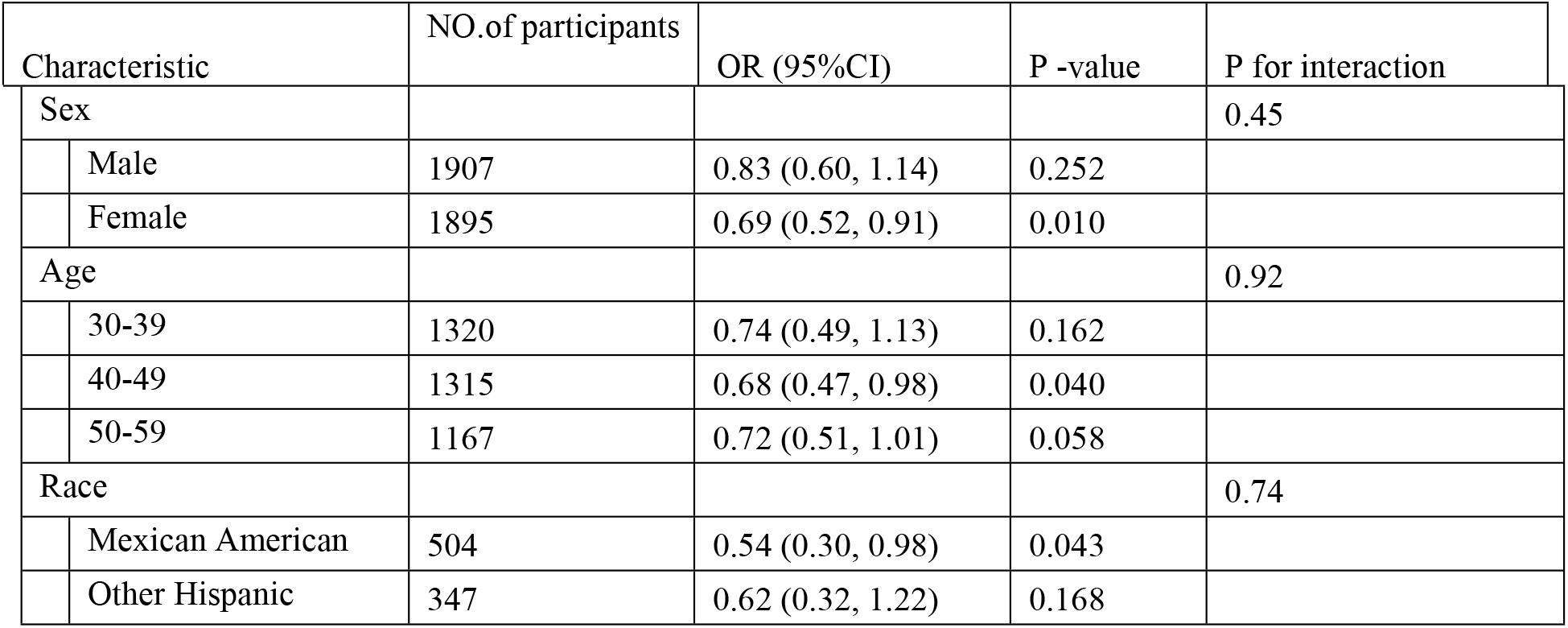
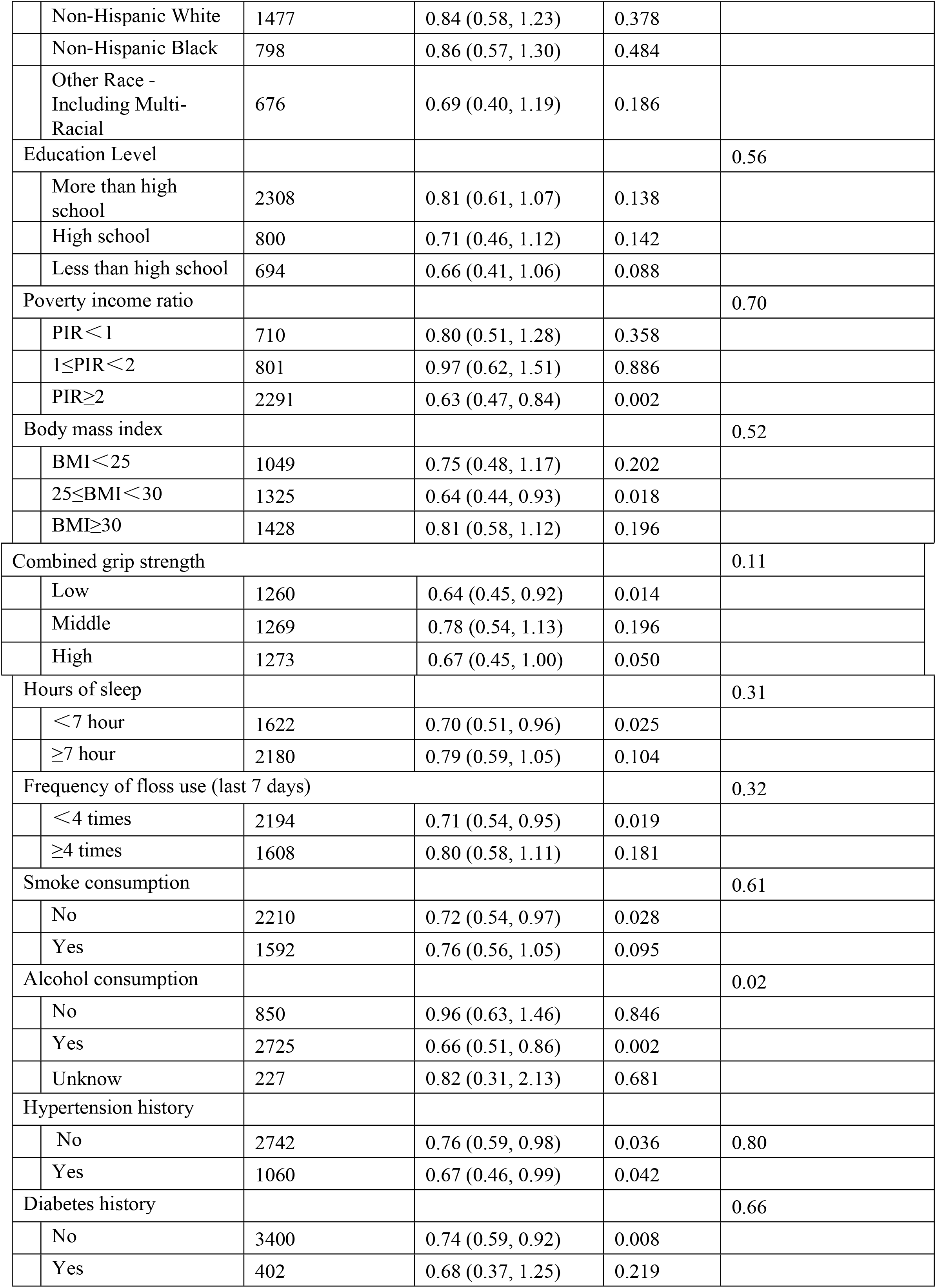
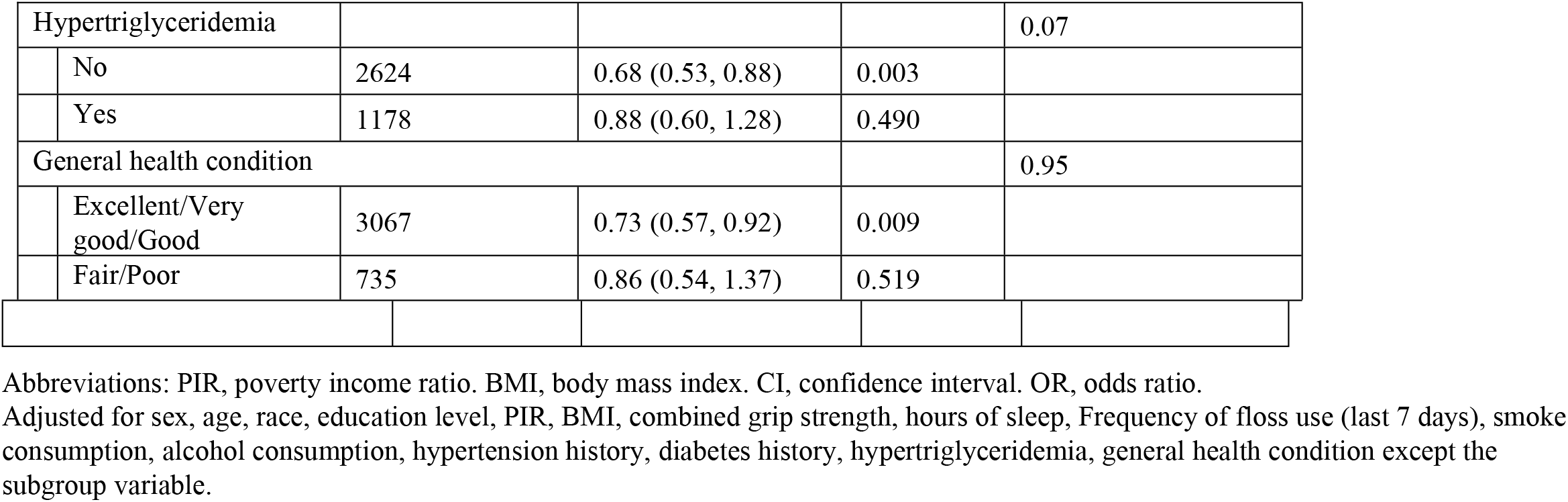
Effect of size of the skull bone mineral density on periodontitis in prespecified and exploratory subgroups in Each Subgroup.

| Characteristic | NO.of participants | OR (95%CI) | P -value | P for interaction |
| --- | --- | --- | --- | --- |
| Sex |  |  |  | 0.45 |
| Male | 1907 | 0.83 (0.60, 1.14) | 0.252 |  |
| Female | 1895 | 0.69 (0.52, 0.91) | 0.010 |  |
| Age |  |  |  | 0.92 |
| 30-39 | 1320 | 0.74 (0.49, 1.13) | 0.162 |  |
| 40-49 | 1315 | 0.68 (0.47, 0.98) | 0.040 |  |
| 50-59 | 1167 | 0.72 (0.51, 1.01) | 0.058 |  |
| Race |  |  |  | 0.74 |
| Mexican American | 504 | 0.54 (0.30, 0.98) | 0.043 |  |
| Other Hispanic | 347 | 0.62 (0.32, 1.22) | 0.168 |  |

|  |  |  |  |  |  |
| --- | --- | --- | --- | --- | --- |
|  | Non-Hispanic White | 1477 | 0.84 (0.58, 1.23) | 0.378 |  |
|  | Non-Hispanic Black | 798 | 0.86 (0.57, 1.30) | 0.484 |  |
|  | Other Race - Including Multi-Racial | 676 | 0.69 (0.40, 1.19) | 0.186 |  |
| Education Level |  |  |  |  | 0.56 |
|  | More than high school | 2308 | 0.81 (0.61, 1.07) | 0.138 |  |
|  | High school | 800 | 0.71 (0.46, 1.12) | 0.142 |  |
|  | Less than high school | 694 | 0.66 (0.41, 1.06) | 0.088 |  |
| Poverty income ratio |  |  |  |  | 0.70 |
|  | PIR<1 | 710 | 0.80 (0.51, 1.28) | 0.358 |  |
|  | 1≤PIR<2 | 801 | 0.97 (0.62, 1.51) | 0.886 |  |
|  | PIR≥2 | 2291 | 0.63 (0.47, 0.84) | 0.002 |  |
| Body mass index |  |  |  |  | 0.52 |
|  | BMI<25 | 1049 | 0.75 (0.48, 1.17) | 0.202 |  |
|  | 25≤BMI<30 | 1325 | 0.64 (0.44, 0.93) | 0.018 |  |
|  | BMI≥30 | 1428 | 0.81 (0.58, 1.12) | 0.196 |  |
| Combined grip strength |  |  |  |  | 0.11 |
|  | Low | 1260 | 0.64 (0.45, 0.92) | 0.014 |  |
|  | Middle | 1269 | 0.78 (0.54, 1.13) | 0.196 |  |
|  | High | 1273 | 0.67 (0.45, 1.00) | 0.050 |  |
| Hours of sleep |  |  |  |  | 0.31 |
|  | <7 hour | 1622 | 0.70 (0.51, 0.96) | 0.025 |  |
|  | ≥7 hour | 2180 | 0.79 (0.59, 1.05) | 0.104 |  |
| Frequency of floss use (last 7 days) |  |  |  |  | 0.32 |
|  | <4 times | 2194 | 0.71 (0.54, 0.95) | 0.019 |  |
|  | ≥4 times | 1608 | 0.80 (0.58, 1.11) | 0.181 |  |
| Smoke consumption |  |  |  |  | 0.61 |
|  | No | 2210 | 0.72 (0.54, 0.97) | 0.028 |  |
|  | Yes | 1592 | 0.76 (0.56, 1.05) | 0.095 |  |
| Alcohol consumption |  |  |  |  | 0.02 |
|  | No | 850 | 0.96 (0.63, 1.46) | 0.846 |  |
|  | Yes | 2725 | 0.66 (0.51, 0.86) | 0.002 |  |
|  | Unknow | 227 | 0.82 (0.31, 2.13) | 0.681 |  |
| Hypertension history |  |  |  |  |  |
|  | No | 2742 | 0.76 (0.59, 0.98) | 0.036 | 0.80 |
|  | Yes | 1060 | 0.67 (0.46, 0.99) | 0.042 |  |
| Diabetes history |  |  |  |  | 0.66 |
|  | No | 3400 | 0.74 (0.59, 0.92) | 0.008 |  |
|  | Yes | 402 | 0.68 (0.37, 1.25) | 0.219 |  |
| Hypertriglyceridemia |  |  |  |  | 0.07 |
|  | No | 2624 | 0.68 (0.53, 0.88) | 0.003 |  |
|  | Yes | 1178 | 0.88 (0.60, 1.28) | 0.490 |  |
| General health condition |  |  |  |  | 0.95 |
|  | Excellent/Very good/Good | 3067 | 0.73 (0.57, 0.92) | 0.009 |  |
|  | Fair/Poor | 735 | 0.86 (0.54, 1.37) | 0.519 |  |

Abbreviations: PIR, poverty income ratio. BMI, body mass index. CI, confidence interval. OR, odds ratio.
Adjusted for sex, age, race, education level, PIR, BMI, combined grip strength, hours of sleep, Frequency of floss use (last 7 days), smoke consumption, alcohol consumption, hypertension history, diabetes history, hypertriglyceridemia, general health condition except the subgroup variable.

### Relationship between skull BMD and periodontitis

The correlation between skull BMD and periodontitis levels is shown in Table 3. When all covariables were considered, the full-adjusted mode indicated that with each additional unit of skull BMD, periodontitis risk decreased by 30% (OR=0.70; CI=0.57, 0.87; p=0.0010), with P for trend lower than 0.05. The skull BMD was stratified by quartile in sensitivity analysis and estimated P for trend to ensure robustness of results. Compared to the reference Q1 group, the estimated increase in skull BMD was 0.91, 0.78, 0.67 respectively for the Q2, Q3, and Q4 groups. The P for trend of 0.0002, the results of skull BMD as a continuous variable were also consistent.

**Table 3.**
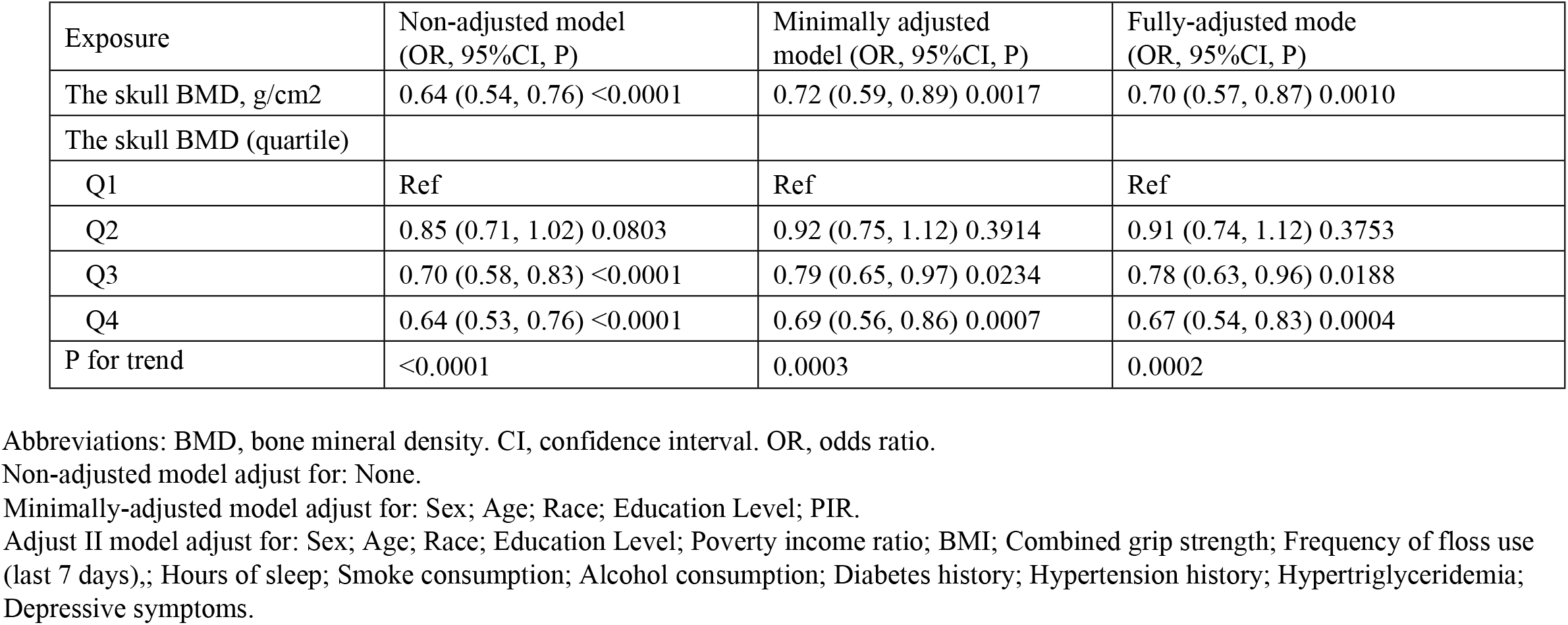
Relationship between skull bone mineral density and periodontitis.

| Exposure | Non-adjusted model<br>(OR, 95%CI, P) | Minimally adjusted<br>model (OR, 95%CI, P) | Fully-adjusted mode<br>(OR, 95%CI, P) |
| --- | --- | --- | --- |
| The skull BMD, g/cm<sup>2</sup> | 0.64 (0.54, 0.76) <0.0001 | 0.72 (0.59, 0.89) 0.0017 | 0.70 (0.57, 0.87) 0.0010 |
| The skull BMD (quartile) |  |  |  |
| Q1 | Ref | Ref | Ref |
| Q2 | 0.85 (0.71, 1.02) 0.0803 | 0.92 (0.75, 1.12) 0.3914 | 0.91 (0.74, 1.12) 0.3753 |
| Q3 | 0.70 (0.58, 0.83) <0.0001 | 0.79 (0.65, 0.97) 0.0234 | 0.78 (0.63, 0.96) 0.0188 |
| Q4 | 0.64 (0.53, 0.76) <0.0001 | 0.69 (0.56, 0.86) 0.0007 | 0.67 (0.54, 0.83) 0.0004 |
| P for trend | <0.0001 | 0.0003 | 0.0002 |

Abbreviations: BMD, bone mineral density. CI, confidence interval. OR, odds ratio.
Non-adjusted model adjust for: None.
Minimally-adjusted model adjust for: Sex; Age; Race; Education Level; PIR. Adjust II model adjust for: Sex; Age; Race; Education Level; Poverty income ratio; BMI; Combined grip strength; Frequency of floss use (last 7 days); Hours of sleep; Smoke consumption; Alcohol consumption; Diabetes history; Hypertension history; Hypertriglyceridemia; Depressive symptoms.

### Dentification of linear relationship

The smooth curve fitting is shown in Fig 1 (Association between skull bone mineral density and periodontitis). We identified a linear correlation between skull BMD and periodontitis using the generalized additive model (Table 4). We compared linear regression and two-piecewise linear regression, the point of inflection was 2.89g/cm^2^. For the left and right inflection points, the odds ratio and confidence intervals are 0.65 (0.50-0.82) and 1.76 (0.49-6.30), respectively. However, the P value for the log-likelihood ratio test is 0.150 (>0.05), indicating that the linear regression model should be used to fit the data.

**Table 4.** Threshold Effect Analysis of the skull bone mineral density and periodontitis using Piece-wise Linear Regression.

| Outcome | Periodontitis (OR, 95%CI, P) |
| --- | --- |
| Fitting by weighted linear regression model | 0.70 (0.57, 0.87) 0.0010 |
| Fitting by weighted two-piecewise linear regression mode |  |
| Inflection point | 2.89 |
| < 2.89 | 0.65 (0.52, 0.82) 0.0003 |
| ≥ 2.89 | 1.76 (0.49, 6.30) 0.3827 |
| Log likelihood ratio test | 0.152 |

Abbreviations: CI, confidence interval. OR, odds ratio. Adjusted for Sex; Age; Race; Education Level; Poverty income ratio; BMI; Combined grip strength; Frequency of floss use (last 7 days); Hours of sleep; Smoke consumption; Alcohol consumption; Diabetes history; Hypertension history; Hypertriglyceridemia; Depressive symptoms.

**Fig 1.**
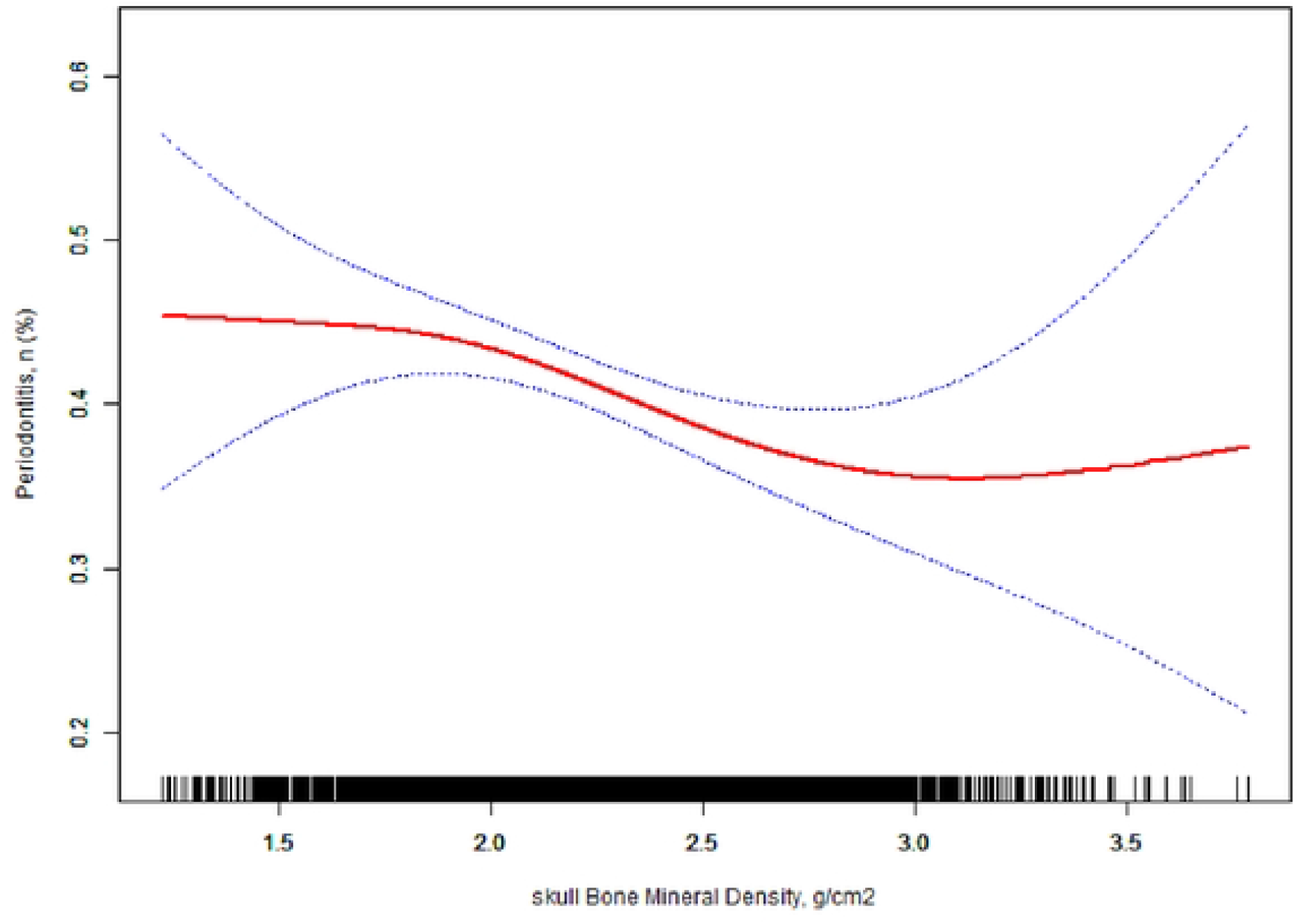
Association between skull bone mineral density and periodontitis. Solid red line represents the smooth curve fit between variables. Blue bands represent the 95% of confidence interval from the fit. All adjusted for Sex; Age; Race; Education Level; Poverty income ratio; BMI; Combined grip strength; Frequency of floss use (last 7 days), Hours of sleep; Smoke consumption; Alcohol consumption; Diabetes history; Hypertension history; Hypertriglyceridemia; Depressive symptoms.

## Discussion

In the present investigation, we revealed that sex, age, race, poverty income ratio, body mass index, hours of sleep, Frequency of floss use (last 7 days), smoke consumption, alcohol consumption, hypertension history, diabetes history, hypertriglyceridemia, general health condition interfere with the relationship between skull BMD and periodontitis. Interestingly, we found that alcohol consumption playing an important role in skull BMD and periodontitis. Drinking is well-known to be an important risk factor for periodontitis(22-24). On the other hand, low-to-moderate alcohol intake has been associated with slowing age-related bone loss by reducing the overall rate of bone remodeling(25). After adjusting for various potential factors, participants with higher skull BMD displayed significantly lower risk of periodontitis, which 1 unit increase in the skull BMD (g/cm^2^) was associated with a 30% (OR=0.70; CI=0.57, 0.87 p=0.0010) reduction in the risk of periodontitis events. The relationship between skull BMD and periodontitis was linear. Although the inflection point was found (the skull BMD= 2.89g/cm^2^), it was not statistically significant. This is, to our knowledge, the first study to utilize a national survey (NHANES) to analyze the association between skull BMD and periodontitis.

Preclinical studies in ovariectomized animals may explain the link between the low skull BMD and periodontitis. Chen et al.(26) performed ovariectomized mice model, which reliably produced osteopenic or osteoporotic phenotype in long bones, successfully constructed the jaw osteoporotic phenotype, 8 weeks later, it was found that the alveolar bone of osteoporotic mice was significantly thinner, along with periodontal ligament(PDL) width and cell density in the PDL were significantly reduced 18% and 25%, respectively, and A reduced number of osteoprogenitor cells, which persisted even after injury, impacted the rate of alveolar bone regeneration. Likewise, Arioka et al.(27) found the same result. Osteoporotic changes in the periodontium seems to create a vicious circle, leading to more severe periodontitis.

Postmenopausal women with osteoporosis are more susceptible to periodontal disease(6-9), which the consequence of estrogen and BMD could be inferred(8). Hunziker et al.(28) suggest that declining estrogen secretion can result in a decrease in mandibular BMD in periodontitis patients. The average age of menopause varies by race and lifestyle, but it is around 50 years in most countries(29). In our study, however, it was not found that the prevalence of periodontitis was higher in females over 50 years old than it was in males. In line with the literature(16), we observed higher skull BMD in woman than in either of the groups of man. The study of Obrant et al.(16) found that women aged 18 to 87 years had significantly higher skull BMD than men of the same age. Wells et al.(30) indicated that women and men have the same bone mineral content in their trunks and limbs before they turn 16, however, from about this age, bone mineral content in all body regions was significantly different between males and females, especially in men, therefore, adult men’s skull bone mass is a small portion of their total bone mass. Paschall et al.(31) investigated the suitability of measuring the skull BMD for estimating age in older adults using DXA and found that a steady increase in the skull BMD in women occurs until they reach 55 years old, at which point an abrupt decline has occurred, in contrast, BMD values in males decrease slightly around 75 years old, but remain steady throughout life. The reason for this difference is the skull BMD did not change synchronously with the rest of the skeleton. Meanwhile, because skull BMD makes up most of the whole body’s BMD, when it is excluded from the whole-body BMD, fracture risk can be better predicted (32, 33).

Furthermore, the mandibular inferior cortex such as mental index (MI) is a part of the panoramic radiograph that can be used to detect osteoporosis in asymptomatic individuals(34, 35). The results of studies indicate that postmenopausal female dental patients who have MI less than 3 mm on panoramic radiographs, osteoporosis or lower skeletal BMD may be a risk(36, 37). Notably, instead of being diagnostic criteria, these clinical indexes tend to warn about osteoporosis. Additionally, it was not found that these index values correlated with either the maxillary or mandibular BMD(38), nor with periodontitis(39).

According to our investigation, data from a large population is used to assess the association between skull BMD and periodontitis for the first time. Moreover, our study included craniofacial bones, used the most ideal method (dual-energy X-ray absorptiometry) to detect BMD. However, despite those advantages, there were a few limitations. Firstly, NHANES data do not include all confounding factors. For example, people with high BMD are not distinguished, because the increase of BMD may also be secondary to a series of potential diseases affecting bone, Secondly, this study can’t be compared with other studies because rarely studied on the skull BMD and periodontitis, the scope of discussion is limited. Therefore, the study could not reflect the most real relationship between skull BMD and periodontitis, and more information should be studied and public health data used.

## Conclusion

Periodontal disease may be related to low skull BMD, for those people, oral hygiene and health care should be more closely monitored.

## Data Availability

The data underlying the results presented in the study are available from the wibsite: https://www.cdc.gov/nchs/nhanes/index.htm.

## Supporting information

**Supplement table 1**. Univariate analysis for periodontitis.

## Acknowledgments

We would like to thank the NHANES databases for making this data available.

## Author Contributions

**Conceptualization**: Fuqian Jin, Jukun Song, Yi Luo

**Formal analysis:** Beichuan Wang, Ming Ding, Jiaxin Hu

**Funding acquisition:** Yi Luo

**Investigation:** Fuqian Jin, Jukun Song, Zhu Chen

**Methodology:** Fuqian Jin, Jiaxin Hu

**Supervision:** Jukun Song, Zhu Chen, Yi Luo

**Writing - original draft:** Fuqian Jin, Beichuan Wang, Ming Ding, Jiaxin Hu

**Writing - review & editing:** Fuqian Jin, Jukun Song, Yi Luo, Beichuan Wang, Ming Ding, Jiaxin Hu, Zhu Chen

## Data availability statement

The NHANES database is available for download

(https://wwwn.cdc.gov/nchs/nhanes/Default.aspx)

## Funding statement

A grant from the Science and Technology Plan Project of Guiyang in 2014([2014]003) and 2019 ([2019]9-7-13).

## Conflict of interest disclosure

All authors declare that they have no conflicts of interest to report.

## Ethics approval statement

Approval of both datasets was granted by NCHS Research Ethics Review Board (ERB), protocol #2011-17 and informed consent was received from all participants.

## Notes

### Competing Interest Statement

The authors have declared no competing interest.

### Funding Statement

The author of YL received the awards from the Science and Technology Plan Project of Guiyang in 2014([2014]003) and 2019 ([2019]9-7-13). URL?http://kjj.guiyang.gov.cn/fzlm/fzlmgywm The funders had no role in study design, data collection and analysis, decision to publish, or preparation of the manuscript.

